# Prevalence and Clinical Impact of Pathogenic Variants in Cardiomyopathy Genes Among Individuals with Cardiac Conduction Disorders

**DOI:** 10.64898/2026.06.13.26355581

**Authors:** Temidayo A. Abe, Favour E. Markson, Quinn S. Wells, Megan C. Lancaster, William G Stevenson, Benjamin M. Shoemaker, Luke Laws, Majd A El-Harasis, Harikrishna Tandri, Travis D Richardson, Jay A Montgomery, Arvindh N Kanagasundram, Dan M. Roden, Giovanni E. Davogustto

## Abstract

**Importance:** Cardiac conduction disorders have traditionally been regarded as a secondary manifestation of underlying structural heart diseases. However, isolated conduction disorders may precede the onset of heart failure (HF) suggesting shared mechanisms.

**Objective:** To evaluate the prevalence and clinical significance of pathogenic/likely pathogenic (P/LP) rare variants in cardiomyopathy genes among individuals with conduction disorders.

**Design, Setting, and Participants:** Biobank analysis of 192,834 participants with whole genome sequence data from Vanderbilt’s BioVU and 353,092 participants from the *All of Us* Research Program (*AoU*). Participants with primary conduction disorder (left bundle branch block [LBBB], right bundle branch block [RBBB], high-grade atrioventricular block [AVB]) were identified after excluding secondary causes.

**Exposures:** P/LP variants in cardiomyopathy genes.

**Main Outcomes and Measures:** Primary outcome was P/LP carrier status by age and HF status. Secondary outcomes included incident HF and composite ventricular arrhythmias/sudden cardiac death/mortality (VA/SCD/mortality).

**Results:** Among 16,959 participants with conduction disorders in BioVU and 13,442 in *AoU*, 432 (2.6%) and 206 (1.5%) were P/LP carriers, respectively. Conduction disorder was independently associated with carrier status (BioVU p<0.001; AoU p=0.005). Carrier probability varied by age at conduction disorder onset and HF status. Among participants with HF at age 30 years, predicted carrier probability for LBBB was 7.5% in BioVU and 20.2% in *AoU*; for high-grade AVB, 7.7% and 8.5%, respectively, compared with 3.7% and 2.9% among those with HF without conduction disorder. P/LP carrier status among participants with conduction disorders was associated with increased risk of incident HF (BioVU p<0.001; *AoU* p<0.001) and ventricular arrhythmia/sudden death/mortality (BioVU p<0.001; *AoU* p<0.001). Carriers also demonstrated increased susceptibility to conduction disorder following HF diagnosis, including more than two-fold higher risk of third-degree AVB (BioVU aOR 2.48, 95% CI 1.85–3.32; *AoU* aOR 2.26, 95% CI 1.35–3.80).

**Conclusions:** Adults with primary conduction disorders have an increased prevalence of P/LP variants in cardiomyopathy genes, which is most pronounced with diagnoses at early ages of adulthood. Furthermore, there is evidence of an interaction between P/LP carrier status and conduction disorder to increase HF risk and composite cardiovascular outcomes, underscoring the potential role of genetic evaluation in patients with primary conduction disorders to inform long-term outcomes.

**Key Points:** *Question:* What is the prevalence of pathogenic/likely pathogenic (P/LP) variants in cardiomyopathy genes among individuals with cardiac conduction disorders, and are carriers at increased risk for adverse cardiovascular outcomes?

*Findings:* In this biobank study of 192,834 participants with whole genome sequence data from Vanderbilt’s BioVU and 353,092 participants from the *All of Us* Research Program, conduction disorder was independently associated with P/LP carrier status, with predicted carrier probability reaching up to 20% among young participants with concurrent heart failure. Carriers with conduction disorder had higher risk of incident heart failure and ventricular arrhythmias, sudden cardiac death, or mortality.

*Meaning:* These findings suggest that genetic testing may be warranted in patients with conduction disorder, particularly younger individuals, and those with heart failure.

## Introduction

Cardiac conduction disorders have traditionally been considered secondary manifestations of underlying structural heart diseases.^1,2^ However, isolated conduction disorders in the absence of manifest structural heart diseases are frequently observed in clinical practice.^3–5^ It is also recognized that isolated cardiac conduction disorders can be early markers of future cardiovascular events, preceding the development of heart failure (HF) by many years.^3–6^

Conduction abnormalities can directly contribute to the development of ventricular dysfunction.^7,8^ Left bundle branch block (LBBB) is present in up to 15% of patients with HF.^9^ LBBB causes left ventricular dyssynchrony that decreases cardiac mechanical efficiency and may drive adverse remodeling, which can cause LBBB-induced cardiomyopathy that can be reversed by cardiac resynchronization therapy.^9–13^ LBBB can, however, be the initial manifestation of an inherited cardiomyopathy due to a pathogenic variant that leads to HF. Thus, LBBB may be causal or concurrent in the pathophysiology of HF.^14,15^

Carriers of pathogenic or likely pathogenic (P/LP) variants in cardiomyopathy genes have greater risk of HF diagnosis, life threatening arrhythmias, and mortality.^16^ Early identification of these patients could potentially guide personalized follow-up and implementation of therapy to attempt to delay or prevent HF and mitigate the risks of sudden death.

With the increasing recognition of the genetic causes of HF, and of dilated cardiomyopathy in particular, the established association between conduction disorders and HF raises the possibility that conduction disorders, including isolated cases, represent the initial phenotypic manifestation of inherited cardiomyopathies.^17,18^ However, the prevalence and clinical relevance of P/LP variants in cardiomyopathy genes among individuals with conduction disorder remain unknown.

We hypothesized that individuals with cardiac conduction disorder have an increased prevalence of P/LP variants in cardiomyopathy genes. We also examined the association of P/LP carrier status with HF and a composite cardiovascular outcome including ventricular arrhythmias, sudden death, and all-cause mortality (VA/SCD/Mortality). We report results from 192,834 participants from Vanderbilt’s BioVU, a large-scale biobank linking whole genome sequencing with longitudinal electronic health records (EHR) and of 353,092 participants from the *All of Us* Research Program *(AoU)*.

## Methods

### Study Oversight

The study protocol was approved by the Vanderbilt University Medical Center (VUMC) Institutional Review Board (VUMC IRB# 251056). Because of the sensitive nature of the data collected for this study, as well as the required privacy regulations of the data source used in this study to safeguard rights of the research participants, requests to access the dataset from qualified researchers trained in human subject confidentiality protocols may be sent to the corresponding author. The *AoU* data are publicly available and accessible to registered researchers.^19^

### Data Source: BioVU

BioVU is the VUMC biorepository that links DNA samples to de-identified EHR. Participants are enrolled through routine clinical care at VUMC, with DNA extracted from discarded blood samples obtained during standard medical procedures, following informed consent.^20,21^ Clinical data were derived from the Synthetic Derivative (SD), a de-identified version of the Vanderbilt University Medical Center’s EHR intended to support research.

We analyzed data from the Alliance for Genomic Discovery (AGD) Cohort, which includes 249,927 BioVU participants who underwent whole-genome sequencing through a partnership among VUMC, Nashville Biosciences, Illumina and Pharma partners. Of these, 141,473 participants had available ECG data, enabling detailed phenotypic characterization of conduction disorders. Additional details about BioVU are available online (https://victr.vumc.org/biovu-description/) and in the Supplemental Material.

### Study Cohort and Outcomes: BioVU

We identified study participants from the AGD cohort using the SD as described in Supplemental Methods. Conduction disorder phenotypes included LBBB, RBBB, and high-grade AVB (any second degree, third-degree, or bi- or tri-fascicular block), identified through the combination of ECG data and diagnostic codes (Supplemental Table 1). Extended phenotypes included nonspecific intraventricular conduction delay (IVCD) and left hemiblocks in supplemental analyses. Participants were categorized by the first conduction disorder phenotype identified in either ECG impression and/or diagnostic codes. We limited case definition to participants with primary conduction disorder by excluding participants with myocardial infarction and coronary revascularization, aortic valve surgery, mitral valve surgery, septal myomectomy, or septal ablation occurring before or within 180 days after the first documented date for each conduction disorder. For controls, the same exclusions were applied without temporal restrictions (Supplemental Figure 1).

The primary outcome was the prevalence of P/LP variants across conduction disorder phenotypes by age. We determined P/LP status in genes with definitive, strong, or moderate evidence for disease association included in curated cardiomyopathy panels from the Clinical Genome Resource (ClinGen) (Supplemental Methods, Supplemental Table 2 and 3). Variants in these genes were categorized as P/LP if they were assessed as such in ClinVar (https://www.ncbi.nlm.nih.gov/clinvar/).

Secondary outcomes included the association of P/LP variant carrier status with incident HF, as well as composite VA/SCD/Mortality stratified by type of conduction disorder. HF was ascertained using a validated algorithm from Phenotype KnowledgeBase (PheKB, https://www.phekb.org/), which captures symptomatic episodes across the spectrum of HF including HF with reduced and preserved ejection fraction (Supplemental Methods, Supplemental Table 1) ^22^.

### Data Source, Study Cohort and Outcomes: All of Us Research Program

We utilized the Controlled Tier Dataset version 8, available through the Researcher Workbench, which included 393,601 participants with available EHR and whole-genome sequencing. Given limited ECG data in *AoU* (approximately 20,000), conduction disorder phenotypes were primarily ascertained using the Observational Medical Outcomes Partnership common data model format. The same exclusion criteria as with BioVU were applied to define primary conduction disorders (Supplemental Figure 2). P/LP variants were identified in the same genes using identical ClinVar classifications. Additional details are provided in the Supplemental Material.

### Statistical Analyses

Statistical analyses were performed with the R statistical program (v4.4.2) and used the rms, survival, dplyr, ggplot2, and gridExtra packages. Unless otherwise specified, continuous variables are presented as median (interquartile range), and categorical variables as counts (percentage). Baseline characteristics were summarized by conduction disorder phenotypes and carrier status. Statistical significance was defined as P<0.05.

In BioVU, all analyses were restricted to subjects receiving ongoing care at our institution, defined as ≥3 encounters within every 5-year period of available medical records to ensure dense phenotyping and to increase confidence in outcomes availability.

To examine the association between conduction disorder phenotypes and P/LP carrier status, we fitted multivariable logistic regression models with P/LP carrier status as the outcome variable. All models were adjusted for sex, and the first 10 principal components of ancestry. For participants with conduction disorder, we used age at conduction disorder onset; for controls without conduction disorder, we used age at first EHR encounter. Age was modeled as a restricted cubic spline with 3 knots (join points) at default locations, which are the 0.1, 0.5, and 0.9 quantiles. These models were stratified by the presence of HF at conduction disorder onset and included an interaction term between age and conduction disorder. To assess the overall association independent of subtype, we performed a non-interaction model pooling all conduction disorder subtypes (any conduction disorder vs no conduction disorder).

For secondary outcomes, we examined associations between P/LP carrier status and incident HF and composite VA/SCD/mortality from first EHR encounter using Cox proportional hazards regression, excluding participants with prevalent disease at first EHR encounter for each analysis. Models compared eight mutually exclusive groups defined by carrier status within each conduction disorder phenotype (no conduction disorder, LBBB, RBBB, high-grade AVB), with non-carriers without conduction disorder as the reference group. A separate model compared any conduction disorder vs no conduction disorder. Covariates included age, sex, hypertension, diabetes, average number of EHR encounters per year, and the first 10 principal components of ancestry. The proportional hazards assumption for Cox models was assessed using Schoenfeld residuals.

## Results

### Baseline characteristics

We identified 16,959 participants with any conduction disorder (LBBB, RBBB, high-grade AVB, or IVCD/hemiblock) in BioVU and 13,442 in *AoU*, of whom 432 (2.6%) and 206 (1.5%) were P/LP carriers, respectively (Supplemental Figures 1 and 2). Baseline characteristics stratified by carrier status are shown in Table 1. Carriers with conduction disorder were younger than non-carriers in both cohorts (median age at conduction disorder onset: BioVU 61.4 [IQR 46.9–70.3] vs 66.6 [55.9–75.0] years; *AoU* 60.4 [47.9–69.6] vs 64.8 [55.0–72.4] years). European ancestry predominated (BioVU: 85%; *AoU*: 57%), and limited sample sizes precluded ancestry-specific variant analyses. There were no major differences in comorbid conditions by carrier status.

**Table 1:**
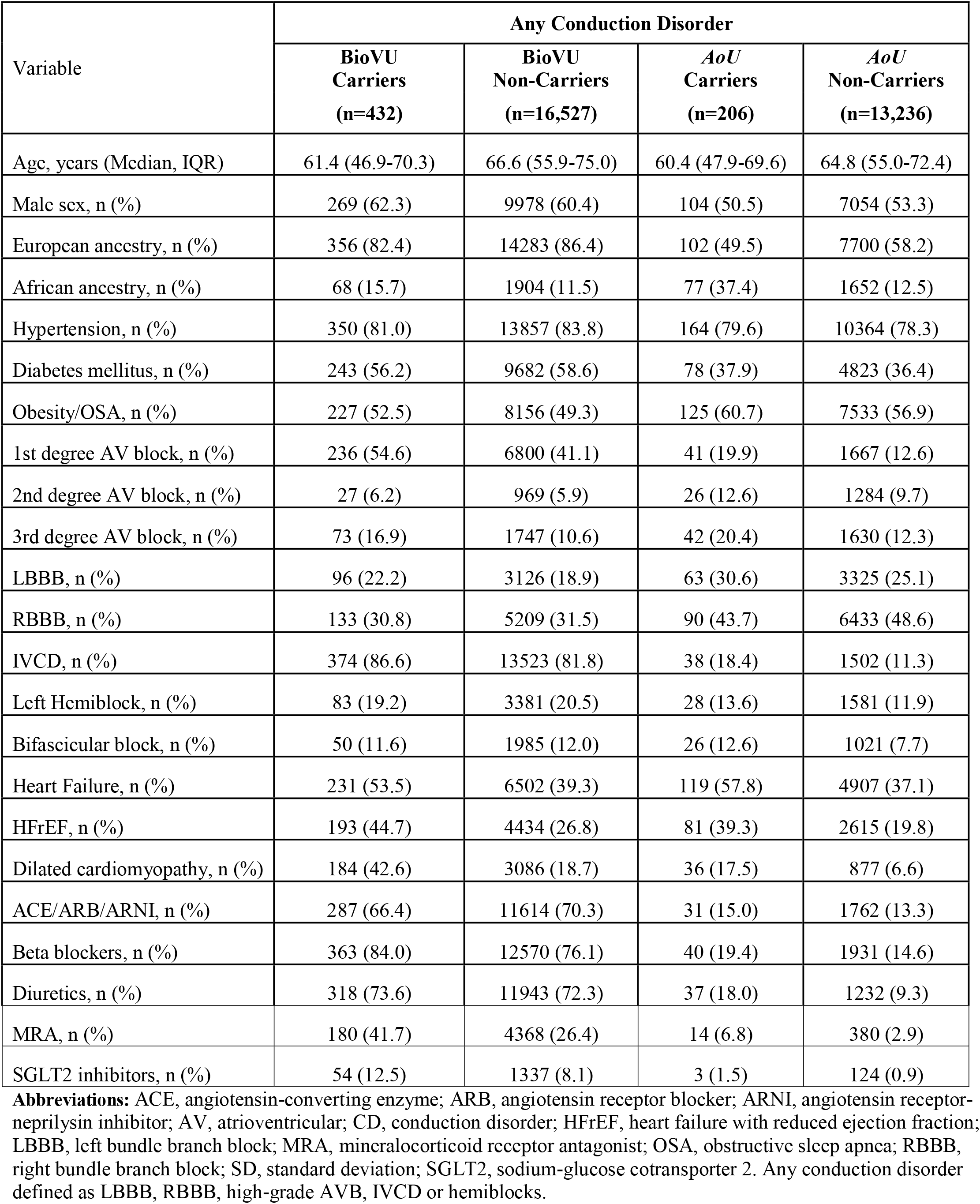
Demographic and Clinical Characteristics by Conduction Disorder and Carrier Status of Pathogenic or Likely Pathogenic variants in Cardiomyopathy Genes.

### Prevalence of Pathogenic/Likely Pathogenic Variants by Conduction Disorder Status

Overall, carrier rates were higher among participants with conduction disorder compared with those without conduction disorder (BioVU: 2.6% vs 1.3%; *AoU*: 1.5% vs 1.3%). Among conduction disorder subtypes, LBBB and high-grade AVB demonstrated the highest carrier rates in both cohorts (BioVU: LBBB 3.1%, high-grade AVB 3.1%; *AoU*: LBBB 1.7%, high-grade AVB 2.2%) (Supplemental Figures 1 and 2).

The presence of conduction disorder and age were independently associated with increased probability of P/LP carrier status in cardiomyopathy genes in both cohorts (conduction disorder, BioVU p<0.001; *AoU* p=0.005; age, BioVU p<0.001; *AoU* p<0.001) (Figure 1). Carrier probability varied by HF status (HF effect, BioVU p<0.001; *AoU* p<0.001), with the strongest differences observed at younger ages (Figure 2A–B). For example, among participants with HF at 30 years of age, carrier probability was 7.5% (95% CI 2.9–16.6%) for LBBB in BioVU and 20.2% (95% CI 6.7–47.1%) in *AoU*. For high-grade AVB, carrier probability was 7.7% (95% CI 3.4–16.4%) and 8.5% (95% CI 2.8–22.9%), respectively, compared with 3.7% (95% CI 2.7–5.1%) and 2.9% (95% CI 2.2–3.7%) among participants with HF but without conduction disorder.

**Figure 1.**
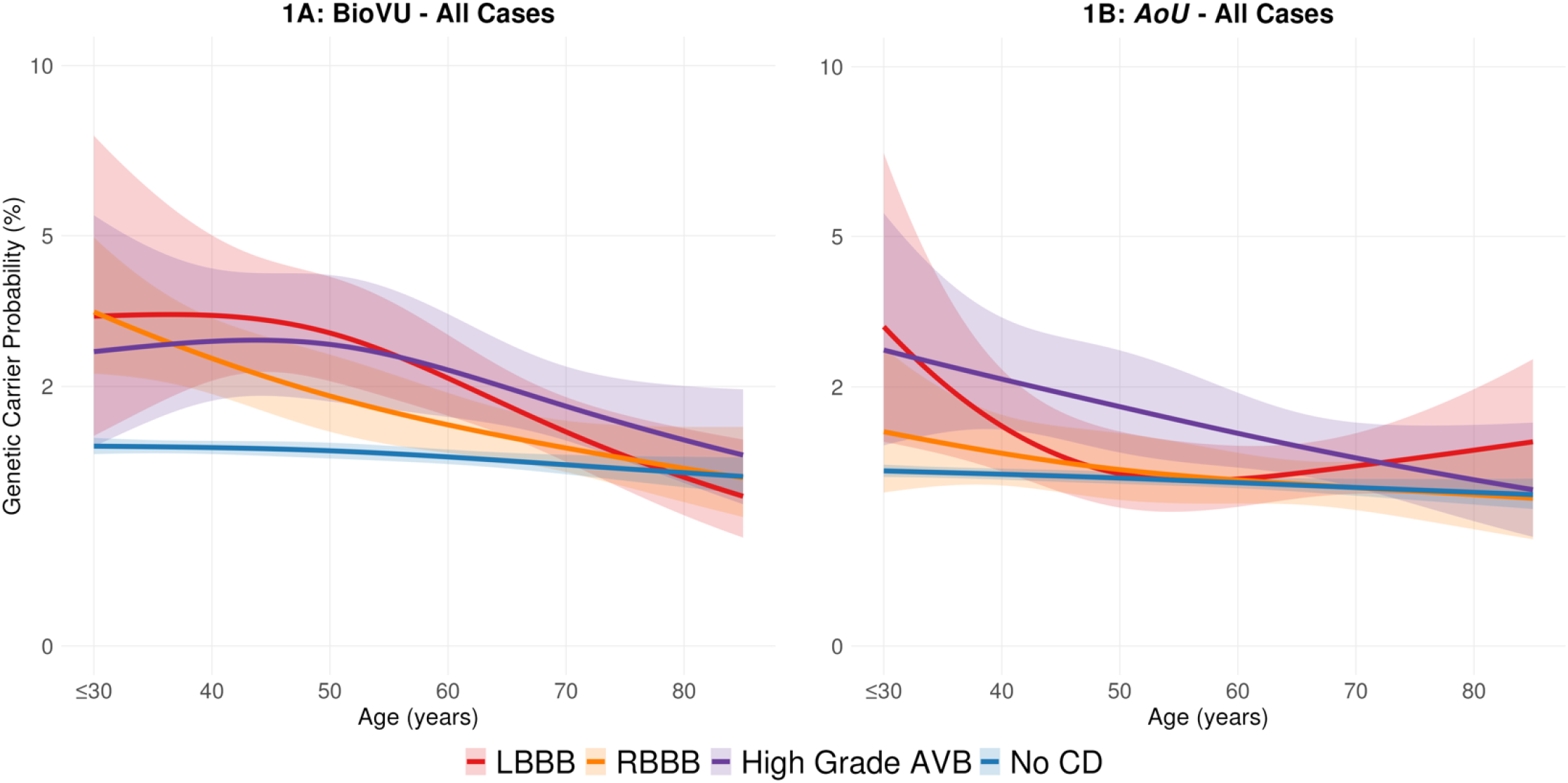
Probability of pathogenic/likely pathogenic carrier status in cardiomyopathy genes by age stratified by type of conduction disorder. Solid lines represent the probability and shaded areas the 95% confidence interval of carrier status for pathogenic/likely pathogenic variants in cardiomyopathy genes by age. Participants were stratified by diagnoses of left bundle branch block (LBBB, red), right bundle branch block (RBBB, orange), high-grade atrioventricular block (purple), and controls without conduction disorder (blue). **(A)** BioVU: conduction disorder (P<0.001) and age (P<0.001) were independently associated with carrier status, with significant age × conduction disorder interaction (P=0.002) and heart failure effect (P<0.001) **(B)** *All of Us (AoU)*: conduction disorder (P<0.001) and age (P<0.001) were associated with carrier status; age × conduction disorder interaction was not significant (P=0.12). Heart failure effect was significant (P<0.001). Models adjusted for sex, heart failure status, and principal components of ancestry 1-10. **Abbreviations:** AVB, atrioventricular block; LBBB, left bundle branch block; P/LP, pathogenic or likely pathogenic; RBBB, right bundle branch block.

**Figure 2.**
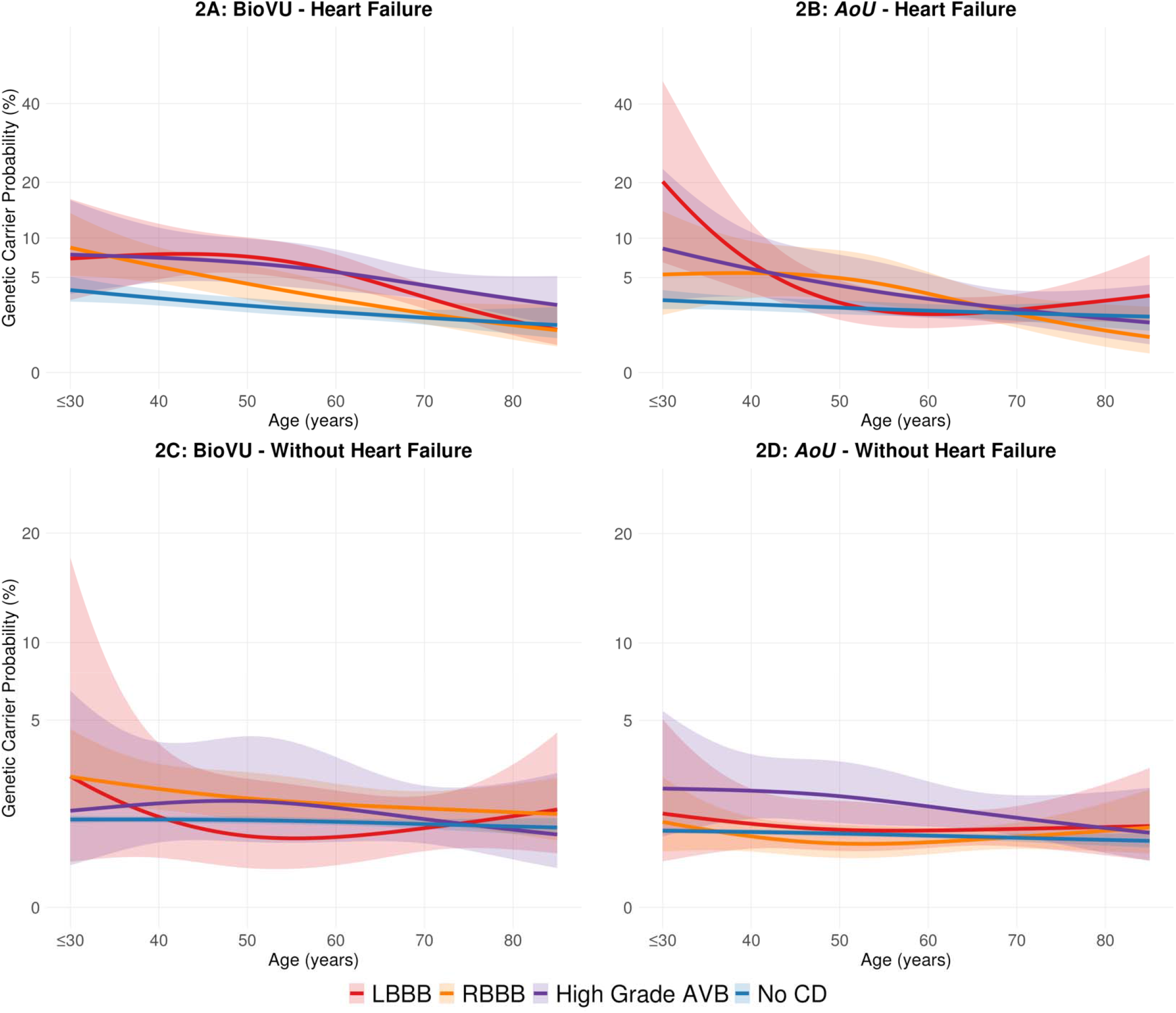
Probability of pathogenic/likely pathogenic carrier status in cardiomyopathy genes by age stratified by type of conduction disorder and heart failure status. Solid lines represent the probability and shaded areas the 95% confidence interval of carrier status for pathogenic/likely pathogenic variants in cardiomyopathy genes by age. Participants were stratified by diagnoses of left bundle branch block (LBBB, red), right bundle branch block (RBBB, orange), high-grade atrioventricular block (purple), and controls without conduction disorder (blue). **(A)** BioVU with heart failure: conduction disorder (P<0.001) and age (P<0.001) were associated with carrier status; age × conduction disorder interaction not significant (P=0.25). **(B)** *All of Us (AoU)* with heart failure: conduction disorder (P<0.001) and age (P<0.001) were associated with carrier status; age × conduction disorder interaction was significant (P=0.02). **(C)** BioVU without heart failure: neither conduction disorder (P=0.10) nor age (P=0.51) was associated with carrier status. **(D)** *AoU* without heart failure: conduction disorder (P=0.04) and age (P=0.04) were associated with carrier status. Models adjusted for sex and principal components of ancestry 1-10. **Abbreviations:** AVB, atrioventricular block; HF, heart failure; LBBB, left bundle branch block; P/LP, pathogenic or likely pathogenic; RBBB, right bundle branch block.

Among participants without HF at conduction disorder onset, conduction disorder remained associated with higher carrier probability in *AoU* (P=0.04) but not in BioVU (P=0.10) (Figure 2C–D), although predicted carrier probabilities were still higher among those with conduction disorder. At age 30, carrier probability for LBBB in BioVU was 2.5% (95% CI 0.3– 17.5%) compared with 1.1% (95% CI 1.0–1.2%) among controls, and for high-grade AVB in *AoU* was 2.0% (95% CI 0.7–5.5%) versus 0.8% (95% CI 0.8–1.0%) among controls. When conduction disorder subtypes were pooled, the presence of any conduction disorder was significantly associated with carrier status in both biobanks and in both patients with and without HF at the time of conduction disorder diagnosis (Supplemental Table 4). Across all phenotypes, carrier probability declined with increasing age, converging to that in the control populations by the eighth decade of life.

Similar age and HF dependent patterns were observed for IVCD and left hemiblock but with smaller effect sizes than other conduction disorder (Supplemental Figure 3). Detailed carrier rates stratified by age, conduction disorder subtype, and HF status are provided in Supplemental Tables 5 and 6.

### Gene Distribution and Gene Specific Conduction Disorder Patterns

Among P/LP carriers, *TTN* and *TTR* were the most common genes with variants, together accounting for approximately 35–45% of carriers across conduction disorder phenotypes (Supplemental Figure 4). Given the high proportion of *TTR* carriers which included common ancestry-specific variants, we performed a sensitivity analysis excluding *TTR* carriers. Similar age- and HF-dependent patterns were observed, with higher adjusted odds of carrier status among those with versus without conduction disorder after excluding *TTR* compared with analyses including *TTR* carriers (Supplemental Figures 5, Supplemental Table 4).

Carriers of *LMNA, TNNT2, DES*, and *FLNC* demonstrated consistently higher prevalence of any conduction disorder than non-carriers (Supplemental Figure 6). Gene-specific patterns varied by conduction disorder subtype and HF status. Among participants with HF, *MYH7* carriers demonstrated higher prevalence of high-grade AVB compared with non-carriers in both cohorts (BioVU 11.6% vs 6.2%; *AoU* 19.1%vs 4.3%). In contrast, among participants without HF at conduction disorder onset, *LMNA* carriers showed marked enrichment for high-grade AVB (BioVU 6.9% vs 0.5%; *AoU* 17.6% vs 0.5%) (Supplemental Figure 7).

### Association of Pathogenic/Likely Pathogenic Carrier Status with Heart Failure and Composite Cardiovascular Outcomes

#### Prevalent Outcomes at EHR Presentation

The prevalence of each outcome by conduction disorder and carrier status is presented in Supplemental Tables 7 and 8. At initial clinical presentation, P/LP carriers demonstrated substantially greater disease burden than non-carriers. HF prevalence was approximately three-to fourfold higher in carriers with and without conduction disorder in BioVU, and among carriers with any conduction disorder and high-grade AVB in *AoU* (Supplemental Table 7). The presence of ventricular arrhythmia at presentation was similarly higher in BioVU, with approximately threefold higher odds among LBBB carriers compared with non-carriers (6.2% vs 2.1%; aOR 2.63, 95% CI 1.01–6.85, P=0.04) (Supplemental Table 8). In *AoU*, ventricular arrhythmia analyses at presentation were limited by sample size.

#### Incident Outcomes During Follow Up

Of the 192,834 participants in BioVU and 353,092 in *AoU*, we excluded those with prevalent HF or ventricular arrhythmias at EHR presentation (BioVU: 4,997 with prevalent HF and 419 with prevalent ventricular arrhythmias; *AoU*: 482 and 195, respectively).

Conduction disorder was associated with increased risk of incident HF, and P/LP carrier status conferred additional risk within each group (Figure 3A–B). During median follow up of 15.7 years in BioVU and 11.0 years in *AoU*, P/LP carrier status was associated with 1.5 to 2.5-fold increased HF risk in both cohorts (Supplemental Table 9), with the exception of RBBB carriers in BioVU (aHR 1.43, 95% CI 0.97-2.09, P=0.07).

**Figure 3.**
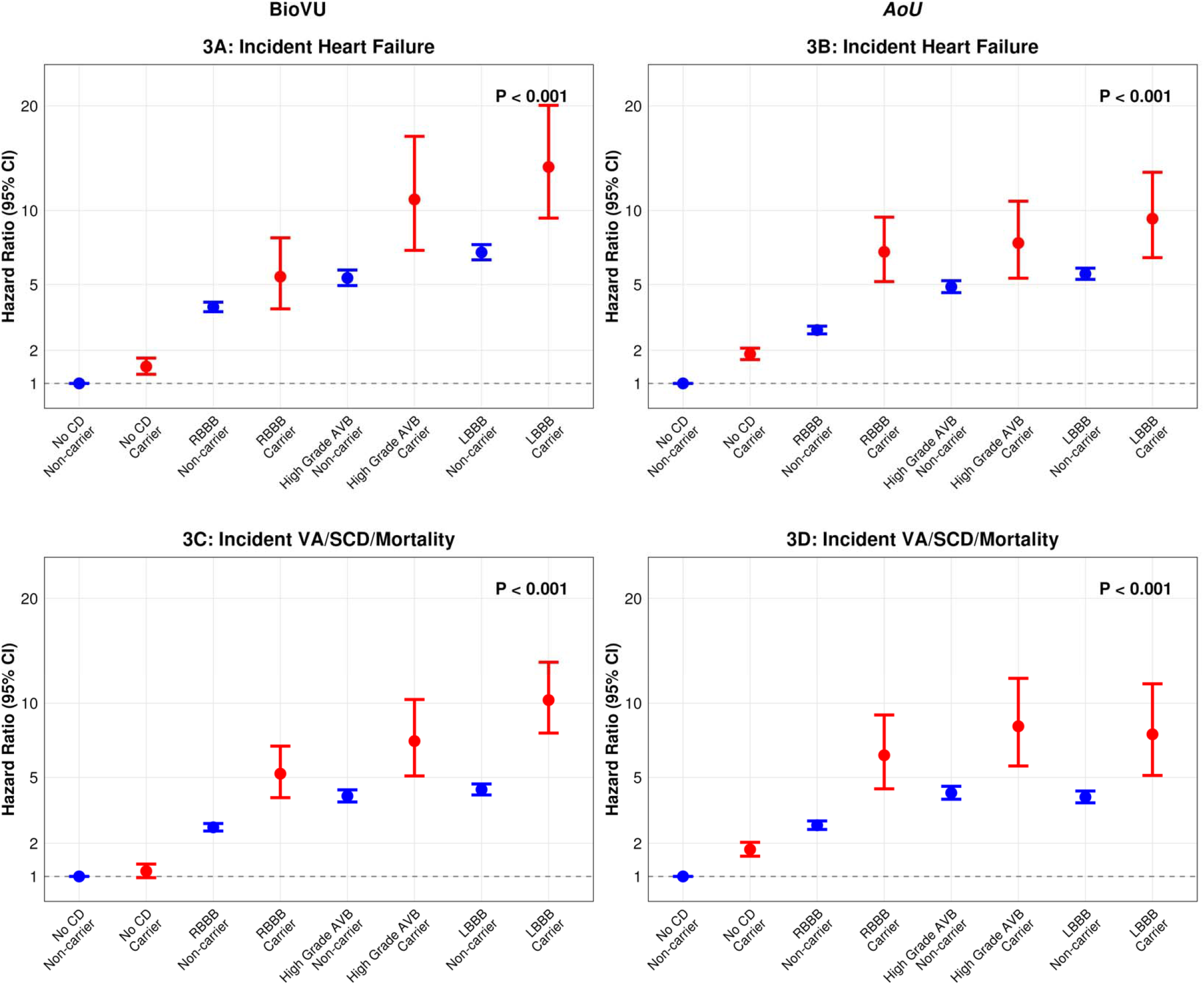
Association of pathogenic/likely pathogenic carrier status in cardiomyopathy genes with heart failure and composite cardiovascular outcome in conduction disorder. Forest plots display hazard ratios with 95% confidence intervals for carriers (red) and non-carriers (blue) compared with non-carriers without conduction disorder (reference, dashed line at 1.0). Models adjusted for age, sex, principal components of ancestry 1–10, hypertension, diabetes, and average number of EHR encounters per year. (A) BioVU incident heart failure. (B) *AoU* incident heart failure. (C) BioVU composite ventricular arrhythmia/sudden cardiac death/mortality. (D) *AoU* composite ventricular arrhythmia/sudden cardiac death/mortality. P-values represent the overall group effect across all conduction disorder phenotypes and carrier status. **Abbreviations:** AVB, atrioventricular block; CD, conduction disorder; LBBB, left bundle branch block; RBBB, right bundle branch block.

During the same follow up time, P/LP carrier status was associated with approximately 2-fold elevated risk across conduction disorder groups for composite VA/SCD/mortality (Figure 3C–D), with consistent directionality for each phenotype across both cohorts (Supplemental Table 10).

### Early and Secondary Conduction Disorder Among Participants with Heart Failure by Carrier Status

Given the significant associations between conduction disorder, P/LP carrier status, and HF, we investigated the temporal relationship between conduction disorder and HF onset in BioVU. Among participants with HF who had ECG data available more than one year before HF diagnosis, P/LP carriers showed higher rates of primary conduction disorder (defined as any first-, second-, or third-degree AVB, RBBB, LBBB, bifascicular block, or QRS duration >120ms) compared with non-carriers although this finding did not reach statistical significance (23.5% (8/34) vs 15.3% (338/2,214); aOR 2.25, 95% CI 0.96–5.27, P=0.061) (Supplemental Figure 8).

Following HF diagnosis, P/LP carriers were more likely to develop new conduction disorders, with nearly 2-fold higher risk for third-degree AVB (BioVU: aOR 2.48, 95% CI 1.85–3.32, P<0.001; *AoU*: aOR 2.26, 95% CI 1.34–3.80, P=0.002) when compared to non-carriers (Supplemental Table 11), particularly among *MYH7* and *MYBPC3* carriers with relatively early (within 5 years) diagnosis of CD after HF diagnosis (Supplemental Figure 8).

## Discussion

In this large-scale analysis involving 545, 926 participants from BioVU and *AoU*, we evaluated the prevalence of P/LP variants in cardiomyopathy genes across conduction disorder phenotypes. We found age-dependent enrichment of P/LP variants among individuals with conduction disorder, most pronounced in the context of HF, where carrier probability for patients under 30 years of age with LBBB reached 20%—up to sixfold higher than controls. This association was attenuated among participants without HF, suggesting HF status as a key modifier of genetic yield. Furthermore, P/LP carriers with conduction disorder had significantly higher risk of incident HF and VA/SCD/mortality compared with non-carriers, with consistent directionality across both cohorts. Importantly, conduction disorder preceded HF diagnosis in both carriers and non-carriers, representing a potential window of opportunity for early genetic identification and intervention.

Although the bidirectional relationship between conduction disorder and HF is well established, the mechanisms linking the two remain poorly understood. This uncertainty has created challenges in management, including variable adherence and response to guideline directed medical therapy, inconsistent benefit from cardiac resynchronization therapy, and overall worse outcomes among patients with HF and conduction disorder.^23–26^ Isolated conduction disorder is particularly difficult to address. Despite its well-documented association with progression to HF, management strategies remain poorly defined. Current guidelines recommend routine monitoring with echocardiography for patients with LBBB and no HF, but optimal surveillance intervals and thresholds for intervention remain unclear.^7^ Furthermore, management of conduction disorder itself adds complexity, as pacing induced cardiomyopathy is frequently but not uniformly observed, suggesting that certain individuals have increased underlying susceptibility.^7,27^

Our findings provide insights into this heterogeneity.^4,28,29^ The enrichment of P/LP variants among younger individuals with conduction disorder, particularly in the context of HF, reinforce the idea that genetic factors may contribute more prominently to conduction disease compared to older ages. Notably, carrier enrichment persisted across conduction disorder phenotypes through presentation age up to 70 years, indicating that the presence of conduction disorder in the setting of HF, even with advancing age, may identify individuals with increased diagnostic yield for genetic testing when secondary causes of conduction disorders have been excluded. Importantly, carriers with conduction disorder demonstrated significantly higher risk of incident HF across all conduction disorder phenotypes, suggesting that genetic testing in patients with conduction disorder may identify individuals at highest risk for progression to cardiomyopathy.^18^

Carriers with conduction disorder similarly demonstrated significantly higher risk of incident VA/SCD/mortality. These associations suggest that conduction disorders may represent an early manifestation of underlying cardiomyopathic processes rather than an isolated conduction system abnormality. Moreover, carriers without conduction disorder at the time of HF onset, demonstrated increased susceptibility to developing conduction disorder after HF diagnosis, with associations for severe phenotypes such as third-degree AVB across both cohorts, supporting the role of genetic susceptibility in both structural and electrical remodeling. This pattern of progressive conduction system involvement throughout the disease course parallels observations described in literature such as with *LMNA*-related cardiomyopathy where conduction disorder is a hallmark feature.^30–33^ Together, these findings highlight the broader consequences of P/LP variants in cardiomyopathy genes and underscore the importance of early genetic testing to identify individuals at risk for HF progression and subsequent conduction disorder, potentially informing surveillance and management strategies.^16,34^

In BioVU participants with HF and ECG data predating HF onset, approximately one in five individuals showed evidence of conduction disorder years before HF, with higher rates among carriers than non-carriers. While our analysis was limited by ECG availability, this observation suggests that conduction disorder may manifest as an early phenotype of cardiomyopathies, preceding overt structural disease by years and representing a potential window for intervention. Importantly, at initial clinical presentation, P/LP carriers already demonstrated substantially greater disease burden than non-carriers, with higher prevalence of HF across conduction disorder phenotypes and ventricular arrhythmias among LBBB carriers. This underscores that by the time patients with conduction disorder present to clinical attention, cardiomyopathic processes may already be well underway, highlighting the importance of early recognition, particularly among carriers of known P/LP variants, where gene-specific management strategies are becoming increasingly available.^35–37^ As precision medicine advances and targeted therapies emerge, early identification of conduction disorder in carriers could enable preventive interventions before irreversible myocardial damage occurs.^38^

### Limitations

Our study has several limitations. Despite combining two large cohorts, the absolute number of P/LP carriers with conduction disorder remained modest, limiting statistical power for gene-specific analyses, and contributing to broad confidence intervals in subgroup analyses. In addition, variant classification was restricted to those already designated as P/LP in ClinVar, likely underestimating true genetic yield by excluding novel, rare, or ancestry-specific variants not yet submitted or classified. Also, we focused exclusively on cardiomyopathy gene panels; inclusion of comprehensive cardiomyopathy-arrhythmia panels would likely increase diagnostic yield further and likely contributes to HF as a key modifier of genetic yield in our study. Similarly, BioVU represents a tertiary referral center population, potentially enriching for more severe phenotypes and limiting generalizability and the cohort was predominantly of European ancestry (~85% of carriers with conduction disorder in BioVU), limiting applicability to diverse populations where variant frequencies and penetrance may differ;^39^ however, *AoU* was more diverse (57% European ancestry) and demonstrated consistent findings. Similarly HF identification using the PheKB algorithm, while validated, requires extensive documentation, and may miss milder phenotypes or early-stage disease. Furthermore, our temporal analysis of early conduction disorder was limited by ECG availability after applying ECG exclusion criteria, precluding definitive conclusions about prevalence. Moreover, variant interpretation was based on current knowledge and may change with reclassification over time. Despite these limitations, our findings provide important insights into the genetic architecture of conduction disorders.

## Conclusion

In this large-scale biobank analysis, we demonstrate age-dependent enrichment of P/LP variants in cardiomyopathy genes among individuals with conduction disorders, with the strongest effects observed in younger patients with concurrent HF, where predicted carrier probability was as high as 20%. Carriers with conduction disorder had higher risk of incident HF and composite VA/SCD/mortality, supporting the role of genetic testing in patients with conduction disorder, particularly younger individuals, and those with concurrent HF, and underscoring the need for prospective studies to guide risk stratification and management.

## Data Availability

All of Us is publicly available

## Notes

**Funding and Acknowledgements:** This work was supported by the American College of Cardiology, Association of Black Cardiologists Merck Research Fellowship Award (T.A.A.). GED receives funding from the American Heart Association (24CDA1271003). Vanderbilt University Medical Center’s BioVU is supported by numerous sources: institutional funding, private agencies, and federal grants. These include the NIH funded Shared Instrumentation Grant S10RR025141; and CTSA grants UL1TR002243, UL1TR000445, and UL1RR024975. Genomic data are also supported by investigator-led projects that include U01HG004798, R01NS032830, RC2GM092618, P50GM115305, U01HG006378, U19HL065962, R01HD074711; and additional funding sources listed at https://victr.vumc.org/biovu-funding/.

### Competing Interest Statement

The authors have declared no competing interest.

